# Central Nervous System Manifestations in COVID-19 Patients: A Systematic Review and Meta-analysis

**DOI:** 10.1101/2020.07.21.20158691

**Authors:** Shahrzad Nazari, Amirhossein Azari Jafari, Seyyedmohammadsadeq Mirmoeeni, Saeid Sadeghian, Mohammad Eghbal Heidari, Siavash Sadeghian, Farhad Asarzadegan, Seyed Mahmoud Puormand, Hamid Ebadi, Davood Fathi, Sahar Dalvand

**Author notes:** **Correspondence:** Sahar Dalvand, Department of Epidemiology and Biostatistics, School of Public Health, Tehran University of Medical Sciences, Tehran, Iran. Address: Tehran University of Medical Sciences, Enqelab Square, Tehran, Iran. **Co-correspond:** Davood Fathi, (**Email:**).

## Abstract

**Background:** At the end of December 2019, a novel respiratory infection, initially reported in China, known as COVID-19 initially reported in China, and later known as COVID-19, led to a global pandemic. Despite many studies reporting respiratory infections as the primary manifestations of this illness, an increasing number of investigations have focused on the central nervous system (CNS) manifestations in COVID-19. In this study, we aimed to evaluate the CNS presentations in COVID-19 patients in an attempt to identify the common CNS features and provide a better overview to tackle this new pandemic.

**Methods:** In this systematic review and meta-analysis, we searched PubMed, Web of Science, Ovid, Embase, Scopus, and Google Scholar. Included studies were publications that reported the CNS features between January 1st, 2020, to April 20th, 2020. The data of selected studies were screened and extracted independently by four reviewers. Extracted data analyzed by using STATA statistical software. The study protocol registered with PROSPERO (CRD42020184456).

**Results:** Of 2353 retrieved studies, we selected 64 studies with 11282 patients after screening. Most of the studies were conducted in China (58 studies). The most common CNS symptom of COVID-19 were Headache (8.69%, 95%CI: 6.76%-10.82%), Dizziness (5.94%, 95%CI: 3.66%-

8.22%), and Impaired consciousness (1.9%, 95%CI: 1%-2.79%).

**Conclusions:** The growing number of studies have reported COVID-19, CNS presentations as remarkable manifestations that happen. Hence, understanding the CNS characteristics of COVID-19 can help us for better diagnosis and ultimately prevention of worse outcomes.

## 1. Introduction

At the end of December 2019, a novel respiratory syndrome, currently known as COVID-19, was reported in Wuhan city, Hubei province, China, and the first sign of this 2019 novel coronavirus infection (2019-nCoV, COVID-19) was pneumonia [1-7]. This new infection rapidly spread worldwide, and an increasing number of infected cases and deaths have been reported globally [8, 9]. Hence, the COVID-19 outbreak was officially considered as a Public Health Emergency of International Concern (PHEIC) by the World Health Organization (WHO) Emergency Committee [10, 11]. Severe acute respiratory syndrome coronavirus 2 (SARS-CoV-2) is a zoonotic pathogen and can transmit from infected animals (such as bats and snakes) to humans eventually leading to epidemics and pandemics through human-to-human transmission [11, 12]. Most cases of COVID-19 have shown respiratory symptoms ranging from cough to dyspnea and respiratory failure as well as the typical signs and symptoms of infection such as fever and fatigue [7, 13-15].

However, a growing number of COVID-19 patients are presenting with different combinations of the central nervous system (CNS) manifestations [16-18]. Several case reports have indicated the presence of various CNS complications, including encephalitis, stroke, meningitis, and encephalopathy in COVID-19 patients [19-22]. Furthermore, a large observational study carried out by Mao et al. show the prevalence of the CNS presentations such as dizziness, headache, impaired consciousness, acute cerebrovascular disease, ataxia, and seizure [16]. Therefore, awareness of the different aspects of the short and long-term effects of this virus on the central nervous system could decently guide scientists. In this systematic review and meta-analysis, we assessed the CNS manifestations in COVID-19 cases.

## 2. Method

### 2-1. Search strategy and selection criteria

We performed this systematic review and meta-analysis based on Preferred Reporting Items for Systematic Reviews and Meta-Analyses (PRISMA) guidelines [23], and our study protocol is submitted to PROSPERO (ID: CRD42020184456). We systematically searched Six databases including Google Scholar, Scopus, PubMed, Web of science, Ovid, and Embase for all published articles from January 1^st^, 2020 until April 20^th^, 2020 using the following Medical Subject Heading terms (MESH terms):

(“Wuhan coronavirus” OR “Wuhan seafood market pneumonia virus” OR “COVID19 virus” OR “COVID-19 virus” OR “coronavirus disease 2019 virus” OR “SARS-CoV-2” OR “SARS2” OR “2019-nCoV” OR “2019 novel coronavirus” OR “2019-nCoV infection” OR “2019 novel coronavirus disease” OR “2019-nCoV disease” OR “coronavirus disease-19” OR “coronavirus disease 2019” OR “2019 novel coronavirus infection” OR “COVID19” OR “COVID-19” OR “severe acute respiratory syndrome coronavirus 2” OR “coronavirus*”) AND (“Manifestation, Neurologic” OR “Neurological Manifestations” OR “Neurologic Manifestation” OR “Neurological Manifestation” OR “Neurologic Symptom” OR “CNS” OR “brain” OR “neuro*” OR “headache” OR “dizziness” OR “ataxia” OR “epilepsy” OR “seizure” OR “migraine*” OR “CSF” OR “Cerebrospinal Fluids” OR “Fluid, Cerebrospinal” OR “Fluids, Cerebrospinal” OR “Cerebro Spinal Fluid” OR “Cerebro Spinal Fluids” OR “Fluid, Cerebro Spinal” OR “Fluids, Cerebro Spinal” OR “Spinal Fluid, Cerebro” OR “Spinal Fluids, Cerebro” OR “stroke” OR “vertigo” OR “consciousness” OR “Impaired consciousness” OR “coma” OR “cerebrovascular disease” OR “acute cerebrovascular disease” OR “encephalitis”) alone or in combination with OR and AND operators.

After removing the duplicated records, articles were screened based on their titles and abstracts by two authors (S.S and M.H) independently. The full texts of eligible publications were examined for inclusion and exclusion criteria (A.AJ, S.M, S.S, and M.H). Observational studies reported at least one of the related CNS symptoms in COVID-19 patients without any language, race, country, and gender limitations included for quantitative synthesis. The preprint studies, interventional studies, systematic reviews, case reports, conferences, commentaries, letters, editorial, author responses, correspondence articles, in vitro, animal studies, children population, articles without full text, or unreliable data were excluded. In addition, the reference list of the eligible studies was searched to prevent missing publication and include all related literature. The data were independently extracted (A.AJ. S.M, S.S, S.S, and M.H), and discrepancies were resolved with discussion and consensus by three independent researchers (SH.N, S.D, and F.A).

### 2-2. Data analysis and quality assessment

The desired data was recorded using an excel spreadsheet form that included the title, first author, year and month of publication, type of study, country, total sample size, the sample size of male and female, study design, demographic characteristics, exposure history, clinical manifestation, CNS symptoms, and any reported comorbidity.We assessed the quality of included studies (A.AJ. S.M, S.S, S.S, and M.H), based on the NIH quality assessment tool for observational cohort and case series studies [24]. This instrument assessed the quality of included studies based on the research question, study population, the participation rate of eligible persons, inclusion and exclusion criteria, sample size justification, analyses, reasonable timeframe, exposure, outcome measures, outcome assessors and, loss to follow-up.

### 2-3. Meta-analysis

Data from included studies were extracted for the number of events and total patients to perform a meta-analysis. Cochrane’s *Q* test and the *I*^2^ index were used to assess heterogeneity among selected studies. Heterogeneity was categorized as low (below 25%), moderate (25%–75%), and high (above 75%) [25]. Also, data adjusted by Freeman-Tukey double arcsine transformation and their 95% CIs were calculated by the Clopper-Pearson method [26]. We calculate mean and standard deviations from median and quartiles by using Wan method [27]. For continuous data, we estimate pooled results of means and their respective 95% CI by the inverse variance method. All analysis were performed using STATA statistical software, version 13 (StataCorp).

## 3. Results

As illustrated in (Figure 1), a total of 2353 studies were retrieved after a systematic search in the aforementioned databases. After removing duplicates, 1760 studies remained. Then, we narrowed the studies to 203 articles by screening with titles and abstracts. In full-text screening, 45 studies with no reliable or useful data, 24 review articles, 41 preprints, 6 case reports, 1 case controls, 4 reports, 4 papers with specific children population, one study with specific pregnant population, and 13 publications such as Commentary, editorial or Correspondence letters were excluded. Finally, 64 studies [5-7, 15, 16, 28-86] including 11282 COVID-19 patients, met our inclusion criteria, and were entered in meta-analysis. The main characteristics of our included studies are presented in (Table 1).

**Figure 1.**
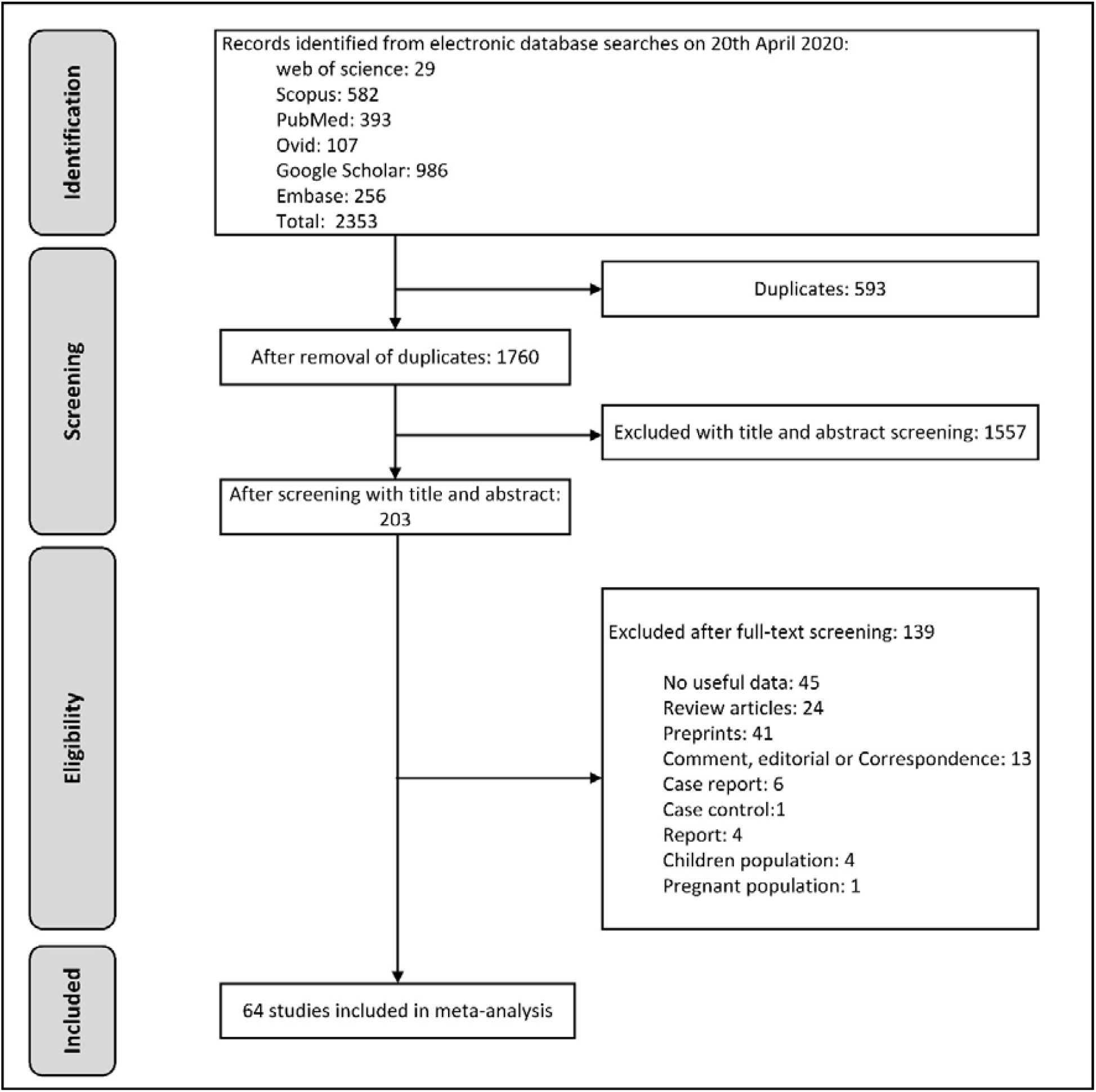
The process of surveying, screening, and selecting the articles for this systematic review and meta-analysis based on PRISMA guideline

**Table 1.** Demographics and baseline characteristics of included studies with COVID-19 infected patients presenting CNS symptoms

| Author | Month of Publication | Type of Studies | Country, City / Province | Sample size | No of Positive Cases (Female/Male) | Quality assessment |
| --- | --- | --- | --- | --- | --- | --- |
| Yang, X [28] | February, 2020 | Cohort | China, Wuhan | 52 | 17 / 35 | Good |
| Yang, W [29] | February, 2020 | Cohort | China, Wenzhou | 149 | 68 / 81 | Fair |
| Wang, D [7] | March, 2020 | Case series | China, Wuhan | 138 | 63 / 75 | Good |
| Song, F [30] | February, 2020 | Retrospective | China, - | 51 | 26 / 25 | Good |
| Shi, H [31] | March, 2020 | Cohort | China, Wuhan | 81 | 39 / 42 | Good |
| Qian, G.-Q [32] | March, 2020 | Case series | China, Zhejiang province | 91 | 54 / 37 | Good |
| Mao, L [16] | April, 2020 | Case series | China, Wuhan | 214 | 127 / 87 | Good |
| Liu, Y [33] | February, 2020 | Case series | China, - | 12 | 4 / 8 | Fair |
| Liu, K [34] | February, 2020 | Retrospective | China, Wuhan | 137 | 76 / 61 | Fair |
| Li, Y [35] | March, 2020 | Cross-Sectional | China, - | 54 | 32 / 22 | Fair |
| Du, Y [36] | April, 2020 | Cohort | China, Wuhan | 85 | 23 / 62 | Fair |
| Cheng, JL [37] | March, 2020 | Cross-Sectional | China, - | 1,079 | 505 / 573 | Fair |
| Chen, N [15] | January, 2020 | Retrospective | China, Wuhan | 99 | 32 / 67 | Good |
| Zhu, W [38] | March, 2020 | Retrospective | China, Anhui province | 32 | 17 / 15 | Good |
| Zhou, F [5] | March, 2020 | Cohort | China, Wuhan | 191 | 72 / 119 | Good |
| Zhong, Q [39] | March, 2020 | Cohort | China, Wuhan | 49 | 42 / 7 | Fair |
| Zheng, Y [40] | April, 2020 | Case series | China, Changsha | 99 | 48 / 51 | Fair |
| Zheng, F [41] | March, 2020 | Retrospective | China, Changsha | 161 | 81 / 80 | Good |
| Zhao, W [42] | March, 2020 | Retrospective Cohort | China, Changsha | 118 | 58 / 60 | Good |
| Zhang, X [43] | March, 2020 | Retrospective | China, - | 573 | 278 / 295 | Good |
| Zhang, J [44] | April, 2020 | Retrospective Cohort | China, Wuhan | 663 | 342/321 | Good |
| Yu, F [45] | March, 2020 | Prospective Cohort | China, Beijing | 76 | 38/38 | Fair |
| Xu, Y. H [46] | February, 2020 | Retrospective | China, Beijing | 50 | 21 / 29 | Fair |
| Wu, J [47] | February, 2020 | Cross- | China, Chongqing | 80 | 38 / 42 | Fair |
|  |  | Sectional |  |  |  |  |
| Wang, X [48] | March, 2020 | Retrospective | China, Wuhan | 1012 | 488 / 524 | Good |
| Wang, L [49] | March, 2020 | Retrospective study | China, Wuhan | 339 | 173 / 166 | Good |
| Wan, S [50] | March, 2020 | Cross sectional | China, Chongqing | 123 | 57 / 66 | Good |
| Wan, Suxin [51] | 2020 | Case series | China, Chongqing | 135 | 63 / 72 | Good |
| Tian, S. J. [52] | February, 2020 | Retrospective observational | China, Beijing | 262 | 135 / 127 | Fair |
| Tan, C [53] | April, 2020 | Retrospective observational | China, Changsha | 27 | 16 / 11 | Fair |
| Sun, C [54] | April, 2020 | Retrospective observational | China, Nanyang | 150 | 83 / 67 | Good |
| Shi, S [6] | March, 2020 | Retrospective observational | China, Wuhan | 416 | 211 / 205 | Fair |
| Shao, F [55] | April, 2020 | Retrospective observational | China, Wuhan | 136 | 46 / 90 | Good |
| Qin, C [56] | March, 2020 | Retrospective observational | China, Wuhan | 452 | 217 / 235 | Good |
| Peng, Y.D [57] | March, 2020 | Retrospective observational | China, Wuhan | 112 | 59 / 53 | Fair |
| Peng, L [58] | April, 2020 | Cross-sectional | China, Shanghai | 86 | 47 / 39 | Fair |
| Moein, S.T [59] | 2020 | Retrospective observational | Iran, Tehran | 60 | 20 / 40 | Fair |
| Mo, P [60] | 2020 | Retrospective | China, Wuhan | 155 | 69 / 86 | Good |
| Mi, B [61] | 2020 | Retrospective | China, Wuhan | 10 | 8 / 2 | Fair |
| Lo, I.L [62] | March, 2020 | Retrospective | China, Macau | 10 | 7 / 3 | Fair |
| Liu, M [63] | February, 2020 | Retrospective | China, Wuhan | 30 | 20 / 10 | Fair |
| Ling, L [64] | 2020 | Retrospective | China, Wuhan | 8 | 4 / 4 | Poor |
| Liang, T [65] | March, 2020 | Retrospective | China, Wuhan | 88 | 37 / 51 | Good |
| Lei, S [66] | 2020 | Retrospective | China, Wuhan | 34 | 20 / 14 | Good |
| Lei, P [67] | 2020 | Retrospective | China, Guiyang | 14 | 6 / 8 | Good |
| Kong, I [68] | February, 2020 | Case series | South Korea, National survey | 28 | 13 / 15 | Poor |
| Kim, E.S [69] | 2020 | Retrospective | Korea, National survey | 28 | 13 / 15 | Good |
| Jin, X [70] | March, 2020 | Retrospective | China, Zhejiang | 651 | 320 / 331 | Good |
| Jia, J [71] | 2020 | Retrospective | China, Qingdao | 44 | 29 / 15 | Fair |
| Huang, C.L [72] | January, 2020 | Retrospective | China, Wuhan | 41 | 11 / 30 | Good |
| Hsih, W.-H [73] | 2020 | Retrospective | Taiwan, Taichung | 43 | 26 / 13 | Fair |
| Han, R [74] | 2020 | Retrospective | China, Wuhan | 108 | 70 / 38 | Fair |
| Gupta, N [75] | April, 2020 | case series | India, New Delhi | 21 | 7 / 14 | Fair |
| Guo, W [76] | 2020 | Retrospective | China, Wuhan | 174 | 98 / 76 | Good |
| Guan, W.-j [77] | Feb, 2020 | cross | China, National | 1099 | 459 / 640 | Good |
| Feng, Y [78] | April, 2020 | Retrospective | China, Wuhan,<br>Shanghai and<br>Anhui | 476 | 205 / 271 | Good |
| Du, R.-H [79] | April, 2020 | Retrospective | China, Wuhan | 109 | 35 / 74 | Good |
| Ding, Q [80] | March, 2020 | case series | China, Wuhan | 5 | 3 / 2 | Good |
| Chen, Y [81] | April, 2020 | Retrospective | China, Wuhan | 42 | 27 / 15 | Fair |
| Chen, X [82] | April, 2020 | Retrospective | China, - | 104 | 52 / 52 | Good |
| Chen, T [83] | March, 2020 | case series | China, Wuhan | 274 | 103 / 171 | Good |
| Chen, J [84] | March, 2020 | Retrospective | China, Shanghai | 249 | 123 / 126 | Good |
| Chen, C [85] | March, 2020 | retrospective | China, Wuhan | 150 | 66 / 84 | Good |
| Barrasa, H [86] | 2020 | case series | Spain, Vitoria | 48 | 21 / 27 | Fair |

The total sample size of eligible studies was 11687, including 5568 females and 6114 males. The mean age for non-critical patients was 48.557 (95% CI: 44.816%-52.299%) and for critical patients was 58.965 (95% CI: 55.792%-62.139%). As shown in (Table 2), the proportion of patients with travel history to Wuhan, Wuhan related exposure, and Living in Wuhan was 30.43%, 3.83%, and 4.74%, respectively. In addition, the proportion of patients with travel history to other infected areas and contact with patients was 1.17% and 13.33%, respectively. Mortality was assessed in 25 studies with a pooled incidence rate of 10.47%. The incidence rate of positive females and males were 46.4% (95% CI: 43%-49.8%) and 49.5% (95% CI: 45.7%-53.3%), respectively. 36.1% (95% CI: 27.9%-44.8%) of infected patients were in the severe, critical, or Intensive care unit condition. In addition, the incidence rate of mortality and survival were 10.47% (95% CI: 5%-17.3%) and 81.43% (95% CI: 65.7%-93.2%), respectively.

**Table 2.** Positive PCR, severity, mortality and exposure history of COVID-19 infected patients having CNS symptoms

| Variable | No of studies | Total sample size | No positive case | Incidence rate (95% CI) | Heterogeneity |  |  |
| --- | --- | --- | --- | --- | --- | --- | --- |
|  |  |  |  |  | I<sup>2</sup> (%) | Q | P-Value |
| Positive Female | 60 | 11425 | 5363 | 0.4642 (0.4301-0.4983) | 92.2 | 752.6 | <0.0001 |
| Positive Male | 60 | 11425 | 5919 | 0.4950 (0.4570-0.5331) | 93.8 | 957.8 | <0.0001 |
| Severe or critical or ICU | 40 | 9821 | 2611 | 0.3617 (0.2791-0.4484) | 98.6 | 2827.4 | <0.0001 |
| Non-Severe or Non-Critical or Non-ICU | 37 | 8095 | 5694 | 0.7061 (0.6229-0.7832) | 98.3 | 2154.0 | <0.0001 |
| Mortality | 25 | 7087 | 556 | 0.1047 (0.0508-0.1733) | 98.4 | 1497.6 | <0.0001 |
| Survival | 18 | 3174 | 2585 | 0.8143 (0.6575-0.9329) | 98.9 | 1600.9 | <0.0001 |
| **Exposure History** |  |  |  |  |  |  |  |
| Travel history to Wuhan | 27 | 6476 | 3434 | 0.5115 (0.3295-0.6920) | 99.5 | 5434.9 | <0.0001 |
| Wuhan related exposure | 2 | 567 | 433 | 0.7852 (0.7501-0.8183) | - | - | - |
| Living in Wuhan | 2 | 1151 | 535 | 0.4746 (0.4455-0.5037) | - | - | - |
| Travel history to other infected areas | 3 | 255 | 133 | 0.5221 (0.4609-0.5832) | 0.0 | 1.1 | 0.5664 |
| Contact with patients | 19 | 4422 | 1504 | 0.3465 (0.2976-0.3953) | 90.1 | 181.2 | <0.0001 |
| Family clustering | 6 | 1182 | 254 | 0.2044 (0.1376-0.2712) | 84.9 | 33.1 | <0.0001 |
| Unknown exposure history | 6 | 472 | 63 | 0.1244 (0.0446-0.2042) | 91.3 | 57.7 | <0.0001 |

Based on the results shown in (Table 3 and Figure 2), the most common manifestations were, fever 79.39% (95% CI: 73.94%-84.37%), cough 54.77% (95%CI: 49.10%-60.38%), fatigue 31.13% (95% CI: 25.33%-37.23%), dyspnea 28.47% (95% CI: 21.49%-35.99%), Chest tightness 23.83% (95% CI: 17.84%-29.82%), and shortness of breath 20.42% (95% CI: 13.28%-28.85%).The highest incidence rate among CNS symptoms of COVID-19 patients was for headache (8.69% with 95% CI: 6.76%-10.82%), followed by Dizziness (5.94%, 95%CI: 3.66%-8.22%),and Impaired consciousness (1.9% with 95% CI: 1%-2.79%).

**Table 3.** Clinical Manifestations in COVID-19 infected patients presenting CNS symptoms

| Variable | No of studies | Total sample size | No of positive case | Incidence rate (95% CI) | Heterogeneity |  |  |
| --- | --- | --- | --- | --- | --- | --- | --- |
|  |  |  |  |  | I<sup>2</sup> (%) | Q | p-Value |
| General symptoms |  |  |  |  |  |  |  |
| Fever | 63 | 11537 | 8723 | 0.7939 (0.7394-0.8437) | 97.7 | 2689.5 | <0.0001 |
| Fatigue | 43 | 8638 | 2454 | 0.3239 (0.2678-0.3800) | 97.8 | 1936.5 | <0.0001 |
| Myalgia (Muscle pain or Muscle injury) | 41 | 7479 | 246 | 0.1395 (0.1169-0.1621) | 88.2 | 338.4 | <0.0001 |
| Nasal congestion | 3 | 2684 | 151 | 0.0554 (0.0428-0.0680) | 51.2 | 4.1 | 0.1290 |
| Rhinorrhea | 15 | 3881 | 163 | 0.0447 (0.0258-0.0676) | 83.9 | 87.2 | <0.0001 |
| Dry cough or Cough | 62 | 11507 | 6047 | 0.5477 (0.4910-0.6038) | 97.0 | 2054.7 | <0.0001 |
| Arthralgia | 3 | 204 | 8 | 0.0243 (0.000-0.0785) | 63.6 | 5.5 | 0.0642 |
| Chill | 11 | 3878 | 512 | 0.1802 (0.0834-0.3021) | 98.4 | 637.1 | <0.0001 |
| GI symptoms <sup>a</sup> | 6 | 1795 | 83 | 0.0501 (0.0148-0.0854) | 87.7 | 40.8 | <0.0001 |
| Nausea | 11 | 1934 | 115 | 0.0595 (0.0387-0.0803) | 69.0 | 32.3 | 0.0004 |
| Vomiting | 11 | 2703 | 97 | 0.0322 (0.0255-0.0397) | 23.6 | 13.1 | 0.2183 |
| Nausea and/or Vomiting | 13 | 3160 | 181 | 0.0518 (0.0337-0.0700) | 79.7 | 59.2 | <0.0001 |
| Anorexia or Inappetence | 17 | 2638 | 588 | 0.2052 (0.1393-0.2711) | 96.8 | 508.7 | <0.0001 |
| Diarrhea | 45 | 8270 | 909 | 0.1030 (0.0832-0.1227) | 91.0 | 489.8 | <0.0001 |
| Abdominal pain | 14 | 3132 | 112 | 0.0345 (0.0205-0.0485) | 74.7 | 51.5 | <0.0001 |
| Chest tightness | 9 | 1857 | 468 | 0.2383 (0.1784-0.2982) | 88.3 | 68.6 | <0.0001 |
| Shortness of breath | 26 | 6538 | 1177 | 0.2042 (0.1328-0.2858) | 98.1 | 1329.0 | <0.0001 |
| Dyspnea | 32 | 4793 | 1255 | 0.2847 (0.2149-0.3599) | 96.4 | 859.7 | <0.0001 |
| Chest pain | 13 | 2490 | 68 | 0.0249 (0.0075-0.0490) | 81.4 | 64.4 | <0.0001 |
| Hemoptysis | 13 | 3518 | 71 | 0.0169 (0.0074-0.0289) | 65.2 | 34.5 | 0.0006 |
| Heart palpitations | 2 | 191 | 13 | 0.0671 (0.0316-0.1026) | - | - | - |
| Pharyngodynia or Throat pain or Pharyngalgia or Throat sore | 41 | 9021 | 888 | 0.0983 (0.0767-0.1219) | 89.9 | 399.9 | <0.0001 |
| Coryza or Sputum production or Expectoration | 30 | 7239 | 1909 | 0.2517 (0.1852-0.3182) | 98.4 | 1791.4 | <0.0001 |
| CNS symptoms |  |  |  |  |  |  |  |
| Headache | 48 | 9782 | 897 | 0.0869 (0.0676-0.1082) | 89.5 | 449.5 | <0.0001 |
| Dizziness | 10 | 2296 | 139 | 0.0594 (0.0366-0.0822) | 81.3 | 48.1 | <0.0001 |
| Headache and/or<br>Dizziness | 5 | 558 | 58 | 0.0978 (0.0733-0.1224) | 6.2 | 4.3 | 0.3711 |
| Impaired consciousness | 2 | 877 | 26 | 0.0190 (0.0100-0.0279) | - | - | - |

**Figure 2.**
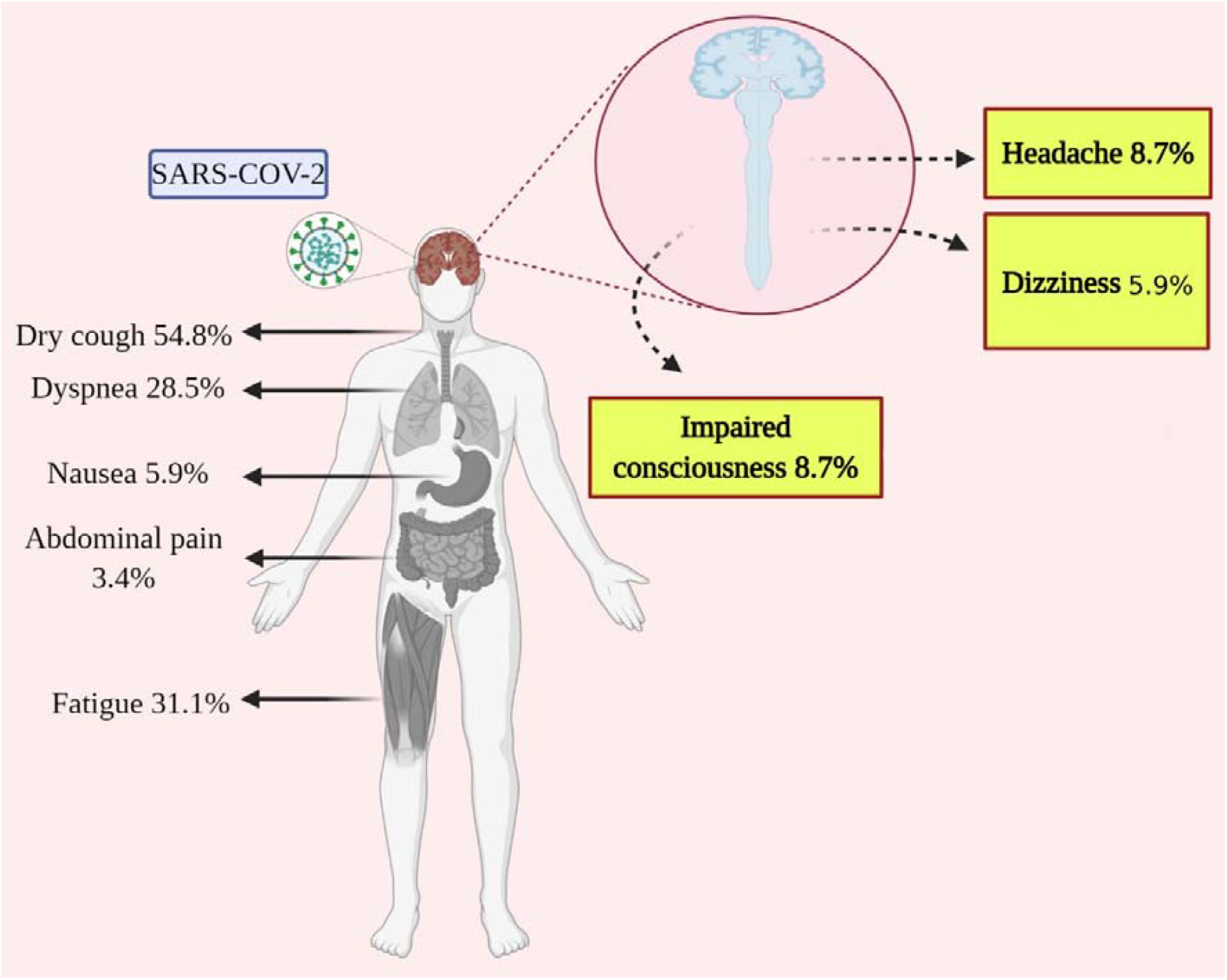
The incidence rate of CNS manifestations in COVID-19 patients (This figure is Created with BioRender.com)

(Table 4) shows comorbidities that were reported in 60 studies including 6959 patients. The highest incidence rate in comorbidities were hypertension with 23.54% (95% CI: 19.14%– 27.94%), diabetes mellitus (11.68% with 95% CI, 0.98%–13.57%), cardiovascular disease (11.66% with 95% CI: 0.89%–14.35%), and cerebrovascular diseases (3.47% with 95% CI:2.29%-4.85%).

**Table 4.**
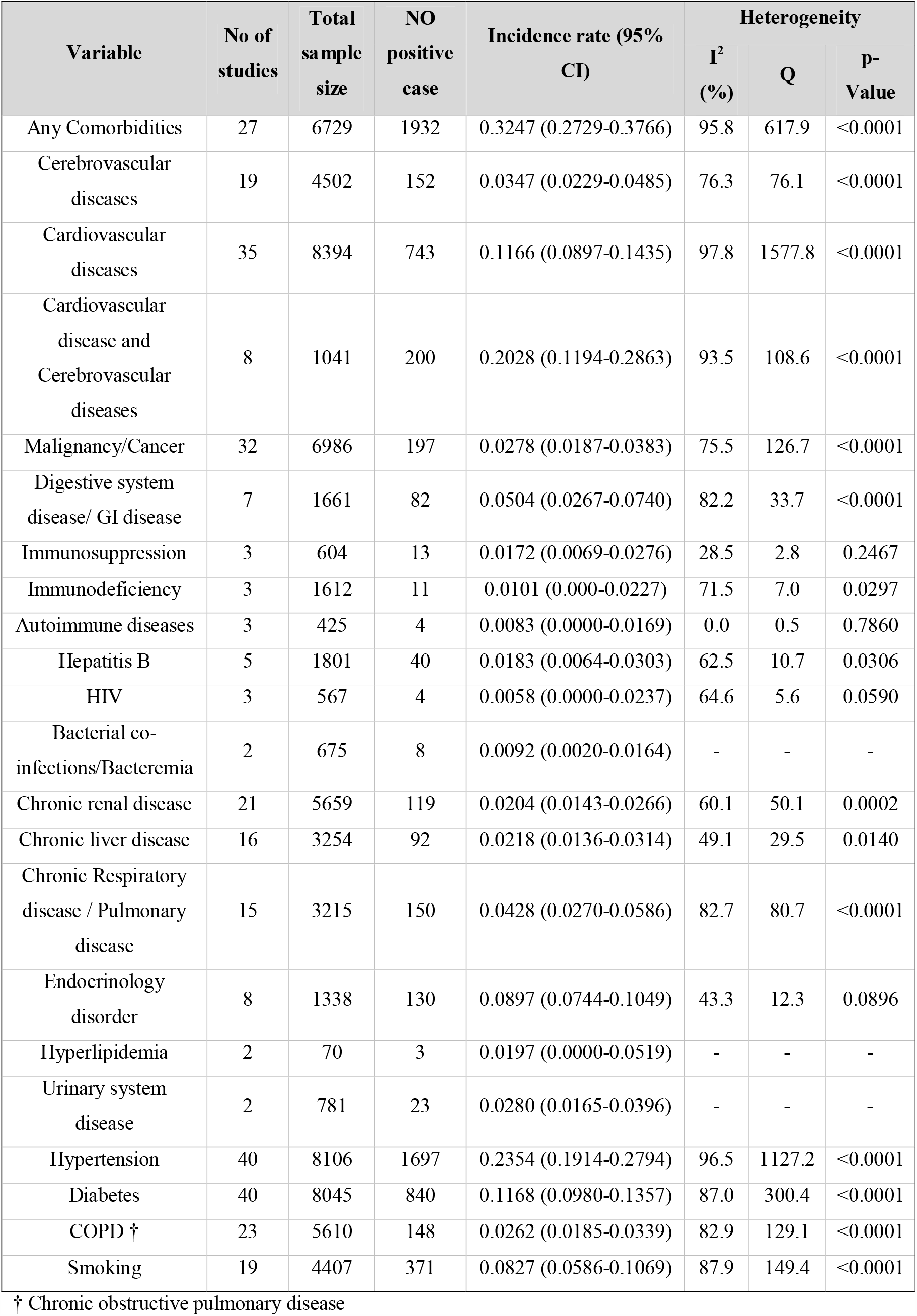
Comorbidities in COVID-19 infected patients with CNS symptoms

## 4. Discussion

Recently, the world has encountered an emergent outbreak posed by the novel coronavirus 2019, officially known as COVID-19 [87]. This infection has become a global threat, endangering millions of lives worldwide [87]. Hence, many experts, researchers, scientists and clinicians are attempting to investigate various aspects of this new infection to find useful solutions for coping with COVID-19 [87]. One of the various aspects of COVID-19 is its impact on the CNS, as reported in a growing number of studies [87]. In addition to the common symptoms in COVID-19, several CNS symptoms such as headache and impaired consciousness have been observed in infected patients [16].

While most investigated the respiratory symptoms of COVID-19, Mao et al. specifically examined the prevalence of neurological manifestations ranging from CNS to peripheral nervous system (PNS) and neuromuscular symptoms in an observational study on COVID-19 patients [16]. They demonstrated CNS presentations ranging from dizziness and headache to impaired consciousness, acute cerebrovascular disease, ataxia, and seizure [16]. Based on the possible neuroinvasive potential of COVID-19, in this systematic review and meta-analysis, we analyzed those evidence indicating the involvement of CNS. We assessed 11687 COVID-19 adult patients from six countries. We reported that COVID-19 patients commonly showed CNS symptoms, including headache, dizziness, and impaired consciousness. Headache (8.69%) were the most common CNS symptoms, followed by dizziness (5.94%) and impaired consciousness (1.9%).

There are two main routes of CNS entry of COVID-19 (hematogenous and peripheral nerves route) leading to CNS infection and inducing various symptoms such as meningitis and encephalitis. In the hematogenous route, the virus infecting respiratory tracts can reach the CNS through the bloodstream via overcoming a strict obstacle known as the blood-brain barrier (BBB) [18, 88-93]. They also may enter the CNS through circumventricular organs, those CNS organs lacking the BBB [94]. The second route, a peripheral nerve, can provide the virus with a retrograde route in to access the CNS via an axonal transport machinery [18, 88-93]. In accordance with this finding, some previous studies on other types of coronaviruses indicating that coronaviruses can reach the brain via cranial nerves (e.g., olfactory, trigeminal nerve terminals in the nasal cavity) [88, 95-97]. Such a neuroinvasive propensity is supported by reporting of patients exhibiting smell impairment, as a hallmark of COVID-19 infection due to the involvement of the olfactory nerve [16, 87].

The COVID-19 infection mechanism requires the virus attaching to its receptor called angiotensin-converting enzyme 2 (ACE2) expressed in various tissues ranging from endothelial cells of the cardiovascular system, airway epithelia, kidney cells, small intestine, and lung parenchyma [98-100]. There exists a wealth of evidence that supports the expression and distribution of the ACE2 in the CNS [100-106]. Hence, ACE2 may be a potential target of COVID-19 upon the entrance into the CNS, triggering its effects on CNS tissue [92]. The presence of the virus in the central nervous system is also supported by some evidence reporting COVID-19 in the CSF of the infected cases [19, 21].

In our meta-analysis, the mortality rate of COVID-19 cases with at least one CNS symptom was 10.47%, which is much higher than the mortality rate of the general infected population [107]. Such a mortality rate can indicate the importance of careful monitoring of CNS manifestations in COVID-19 patients. Moreover, in Mao’s study, the prevalence of CNS manifestations were higher in severe cases of the illness associated with higher mortality [16]. Although, this can be due to the effect of COVID-19 on the brain stem and suppression of the cardiorespiratory control centers causing respiratory failure and death [108], other possible reasons such as cytokine storm and hypoxia have been suggested to explain the high mortality rate in cases with CNS manifestations. Cytokine storm as an immune system response during COVID-19 infection could enhance the permeability of the blood brain-barrier (BBB) [109, 110]. Infection of airway tissues by COVID-19 in severe infection leads to impaired gas exchange, subsequently causing CNS hypoxia resulting in neural dysfunction [111]. This hypoxic condition associated with severe infection disrupts the BBB through elevation of some factors like nitric oxide (NO) and inflammatory cytokines [112]. More precisely, all of these factors mentioned above may contribute to making the BBB more permeable to the virus. Therefore, in the severe condition of infection, COVID-19 easily enters the CNS via disrupted BBB and puts the brain at risk leading to the manifestation of CNS features [113-116].

Furthermore, recent studies have shown that COVID-19 can accelerate the formation of the blood clot in the blood vessels, increasing the risk of cerebrovascular diseases in COVID-19 patients [117, 118]. Hence, because the brain is nourished by a network of blood vessels, this could be indicative of the importance of cerebral vasculature investigations on the CNS symptoms in the COVID-19 infection.

In a nutshell, attention to the CNS aspects of COVID-19 infection has outstanding benefits for clinician’s understanding of a very serious complication of this infection. At this point in time, researchers have mainly focused on finding medicinal treatments for respiratory symptoms of COVID-19. However, it is necessary to investigate the various CNS manifestations of COVID-19 since they are associated with increased severity and mortality [16]. Not only respiratory system dysfunction, but also impairment of respiratory control centers in the CNS (brain stem) can induce acute respiratory failure [108, 119]. Therefore, considering all effective factors, it can provide clinicians to choose the best way in an attempt to manage this pandemic more efficiently.

## 5. Limitations

There are several limitations in our systematic review and meta-analysis. Since in this ongoing pandemic, most of the investigations have conducted on typical signs and symptoms of COVID-19. Thus the number of studies on the atypical complications of COVID-19, such as CNS presentations are partially low. Moreover, there exist many COVID-19 preprint papers that have not yet undergone peer review. Additionally, five studies included in our meta-analysis reported headache and/or dizziness as one symptom in COVID-19 cases. Because we were not sure that headache and/or dizziness is resulted from headache or is a consequence of the dizziness, it would be challenging to categorize headache and/or dizziness in the subgroup of dizziness or headache. Hence, in our meta-analysis, it was not reported as a CNS manifestation and are implied as a separate symptom (table 3).

## 6. Conclusion

COVID-19 is a global problem that currently affects millions of people. This highly pathogenic virus can affect various parts of the human body. Although the respiratory tract has been mainly targeted by COVID-19, the central nervous system can be affected significantly. In addition, patients with more severe illness showed more CNS symptoms, which may bring on worsen clinical conditions. This study achieved an important estimation for the incidence of neurological manifestations in patients with COVID-19. The results of our survey may be helpful for clinicians for better diagnosis and management of CNS signs and symptoms in patients with COVID-19.

## Data Availability

All the related data is available in the manuscript.

## Author Contributors

Study conception and design: SH.N, SD; Acquisition of data: A.AJ. S.M, S.S, S.S, and M.H; Analysis and interpretation of data: SD; Drafting of manuscript: SH.N, SD, A.AJ. S.M, S.S and SM.P; Critical revision: SH.N, SD, A.AJ, F.A, H,E and D.F.

## Acknowledgments

Dr. Katayoun Alikhani for her supports and comments.

